# The effect of the COVID-19 induced lockdown on nutrition, health and lifestyle patterns among adults in Zimbabwe

**DOI:** 10.1101/2020.06.16.20130278

**Authors:** Tonderayi M Matsungo, Prosper Chopera

**Author notes:** Corresponding Author: Dr TM Matsungo.

## Abstract

**Background:** The 2019 coronavirus disease (COVID-19) is a global public health emergency resulting in lockdowns, associated diet and lifestyle changes and constraint public health delivery.

**Objective:** To investigate the impacts of the COVID-19 induced lockdown in Zimbabwe on nutrition, physical activity, alcohol consumption and smoking among Zimbabwean population aged ≥18years.

**Methods:** A cross-sectional online survey was conducted using a structured questionnaire to collect information on demographics (age, gender, place of residence, current employment), food system dimensions, diet and physical activity patterns, stress and anxiety, body image perceptions, lifestyle behaviours like smoking, alcohol intake, screen time, and ease of access to health services. The study obtained ethical clearance from the Medical Research Council of Zimbabwe (MRCZ/B/1920).

**Results:** The participants (n=507) were mostly female (63.0%) between the ages of 31-40 years (48.1%) and had tertiary education (91.3%). The lockdown resulted in increase in food prices (94.8%) and decrease in availability of nutritious foods (64%). Most (62.5%) of the participants reported a reduction in their physical activity levels. The prevalence of Generalised Anxiety Disorder (GAD) was 40.4% and mostly affecting females [63.5%, P=0.909), 31-40 years age group (49.6%, P=0.886). Based on the BMI-based Silhouette Matching Test (BMI-SMT) 44.5% gained weight, 24.3% lost weight and 31.2% did not have weight change. The paired samples T test showed that there was a significant increase in perceived body weight (P<0.001). More than half (59.6%) reported having difficulties accessing medicinal drugs and 37.8% growth monitoring services.

**Conclusions:** The lockdown period was associated with increase in food prices, decrease in dietary diversification, elevated stress, disrupted diet and consumption patterns. There were low levels of physical activity and perceived weight gained during the lockdown period, thus increasing the risk of overweight and obesity. Further studies incorporating participants of different socio-economic status are warranted to get more conclusive results.

**What this paper adds?:**

- First diet and lifestyle survey in Zimbabwe documenting negative effects of lockdown on the urban elite on diets and lifestyles;
- The COVID-19 induced lockdown was associated with elevated anxiety, disruptions of food supply chains and consumption patterns;
- Most of the participants were less active and gained weight in the lockdown period, thus increasing the risk of overweight and obesity an emerging risk factor for severe COVID-19 complications.

## INTRODUCTION

The World Health Organization (WHO) on March 11, 2020 declared the outbreak of the novel 2019 coronavirus disease (COVID-19) a global pandemic ^1^. The pandemic started in Wuhan, China, in late December 2019, and has spread globally ^2^. The WHO situation report 113, reported that as of 12 May 2020 globally there were 4 088 848 cases and 283 153 deaths due to COVID-19 ^3^. The situation report, also showed that the highest cases were in the Americas (1 743 717) and Europe (1 755 790), while Africa had the lowest (46 829).

In order to contain the spread of the virus, national lockdowns characterised by restricted movement and social distancing *“Stay at Home”* have been the order of the day in many countries ^4,5^. In Zimbabwe the government declared a state of national disaster in response to the COVID 19 pandemic on Friday 27 March 2020, this was followed by a nationwide lockdown on March 30 ^6^. Unfortunately this has also led to increased vulnerability to food insecurity especially among the urban poor and increased risk of overweight and obesity in the higher income classes ^7^. This is worrying considering that obesity is a risk factor for severe COVID-19 infection complications ^8^.

Therefore, maintaining healthy body weight and consumption of diverse and nutritious diet is recommended to mitigate COVID-19 infection via the immune boosting mechanism ^9,10^. Individuals who eat a well-balanced diet tend to be healthier with lower risk of obesity and associated complications. In addition, they will have stronger immune systems and reduced risk of contracting infectious diseases like COVID-19 ^11,12^.

Although, the lockdown restrictions have their benefits in “flattening the curve” they also have potential downsides such as increased stress, reduced physical activity, limited availability and access to diverse nutritious foods and health services ^13,14^. In addition, lockdown related stress and anxiety has potential to trigger compensatory hyperphagia “carbohydrate cravings” resulting in increased energy intake thus giving rise to a dangerous vicious cycle ^11,15^. Unfortunately, this food craving is often associated with the increased risk of developing obesity and cardiovascular diseases and has also been linked with increased risk of more serious complications of COVID-19 ^16,17^.

The COVID-19 induced lockdowns are normally characterised with restricted movements and disrupted food supply and accessibility in most settings. Specifically, the COVID-19 control measures and travel restrictions may compromise the ability to maintain physical activity and healthy lifestyles including reduced consumption of diverse and nutritious diets. Although not clearly understood, this can have implications on health and nutrition outcomes. Therefore, this study was designed to investigate the impacts of the COVID-19 induced lockdown in Zimbabwe on nutrition, physical activity and lifestyle patterns among adults in Zimbabwe.

## METHODS

### Study design, setting and participants

The nationwide descriptive cross-sectional study was carried by a nutrition team from the University of Zimbabwe, using a web-survey (SurveyMonkey, California, USA), from 11-25 May 2020. A short online questionnaire was administered with mostly multiple choice questions and some open end questions to capture additional observations. This was a rapid appraisal of the perceptions on COVID-19 lockdown on nutrition, health and lifestyle indicators among adults (≥ 18 years) from the 10 provinces of Zimbabwe (n=507). The survey link was disseminated through institutional and private social networks (WhatsApp, Twitter, Facebook), and institutional mailing lists.

### Data collection and tools

A self-administered online questionnaire was developed to collect data on demographics, socio-economic factors, and to explore the impact of COVID-19 induced lockdown restrictions on food system dimensions, diet and physical activity patterns, stress and anxiety, body image perceptions, lifestyle behaviours like smoking, alcohol intake, screen time (TV/ Tablet/Phone/ Laptop), and ease of access to health services. The survey was carried out using an online platform, which can be accessed through any device with an Internet connection. The survey was disseminated through partner organisations’ mailing lists, institutional and private social media networks (WhatsApp, Twitter, Facebook and LinkedIn).

### Physical activity and lifestyle changes

The perceptions of participants on the impact of COVID-19 on physical activity, screen time, smoking, and alcohol consumption was assessed by asking the question: *What do you perceive as the impact of the Covid-19 induced lockdown on your physical activity /screen time (TV/ tablet/phone/ laptop) / smoking / alcohol consumption?* The responses were coded as follows: 1 =less/ decreased, 2 =same/did not change, 3 =more/increased and 4 =Not applicable.

### Lockdown and food access

In the context of this study food referred to basic everyday foods that households buy from retail outlets and supermarkets. Foods from the basic food groups also known as ‘everyday foods’ provide the nutrients essential for life and growth. Regarding food prices, we were interested in the generic and broader perceptions of participants on the impact of COVID-19 lockdown of prices of basic everyday foods. This important considering that food price hikes can have a strong impact on food affordability, hunger and undernourishment and dietary diversification.

### Diversified diets

The online questionnaire had the following questions to assess perceptions of participants regarding the impact of lockdown on their consumption patterns. (1) Has your diet or consumption patterns changed in the last 4 weeks of lockdown? The next set of questions assessed the impact of the Covid-19 induced lockdown (past 4 weeks) on the usual consumption of the following food groups selected based on the FAO *food groups for assessing household and individual dietary diversity* ^18^ dark green leafy vegetables, other vitamin A rich fruits and vegetables, other vegetables, other fruits, meat and meat groups, cereal breads and tubers, pulses, legumes etc., nuts and seeds, dairy products, eggs ^19^. The responses specific to each food group were coded as follows: 1 =less/ decreased, 2 =same/did not change, 3 =more/increased and 4 =Not applicable.

### Generalized anxiety disorder (GAD)

We used the validated Generalized Anxiety Disorder-7 (GAD-7) scale to assess respondent’s anxiety symptoms ^20^. Seven questions assessed the frequency of anxiety symptoms over the past two weeks on a 4-point Likert-scale ranging from 0 (never) to 3 (nearly every day) and the total score of GAD-7 ranged from 0 to 21, with increasing scores indicated a more severe functional impairments as a result of anxiety ^20^. We adopted the cut-off (GAD total score of >9 points) previously used to define the presence of anxiety symptoms ^21^.

### Body image perceptions

The perceptions of before lockdown and current body images (4 weeks later) of the participants were assessed using the 9 figural BMI-based Silhouette Matching Test (BMI-SMT) ^22^. The silhouettes are ranked from the thinnest to the heaviest body size and responses are recorded on a scale from 1 (thinnest) to 9 (heaviest). The Silhouette test was validated for use in populations of African descent ^22,23^. To assess respondents perceived ideal body size, participants were asked to select a matching figure that best represents how they looked like before the lockdown and how they were looking like the time of the interview. The BMI-SMT silhouette figures are equal in height and increase proportionally in size and are gender specific. In addition, the BMI-SMT provides a reliable and pragmatic method for quantifying body-image perceptions ^22^.

### Data analysis

Data collected was entered and analysed using SPSS v 20 (IBM Inc. Armonk, NY). The Shapiro-Wilk test and data visualisations via Q-Q plots were used to test for normality of data. Continuous data was presented as mean ± standard deviation (SD) while categorical data was presented as frequencies and percentages. Pearson’s Chi-square was used to explore associations for categorical variables and continuous variables respectively. Paired samples T test was used to test for difference in means across continuous normally distributed variables. Level of significance was set at P<0.05.

### Ethics

The study was conducted based on the ethical principles of respect, justice and confidentiality summarised in the 2013 Declaration of Helsinki ^24^. The study obtained ethical clearance from the Medical Research Council of Zimbabwe (MRCZ/B/1920). Electronic informed consent was obtained from all participants prior to completing the survey.

## RESULTS

### Characteristics of participants

On the 25^th^ of May 2020, the online survey was stopped, and the collected data was analysed. In total 507 participants took part in the survey after 2 declined to participate. Most of the participants were between the ages of 31-40 years, were female (63.0%) and had tertiary education (91.3%). Most of the participants were employed in the formal sector (73.9%), and most were from Harare (60.5%). Almost all the participants (95.5%) said they did not receive government assistance during the lockdown period. Of those who did receive some form of assistance, 2.5% received food handouts and 2% received financial assistance (**Table 1**).

**Table 1:** Sociodemographic characteristics of the participants

| Variable | n | % |
| --- | --- | --- |
| *Age category (years)* |  |  |
| 18-30 | 117 | 26.0 |
| 31-40 | 216 | 48.1 |
| 41-49 | 69 | 15.4 |
| 50+ | 47 | 10.5 |
| *Gender* |  |  |
| Male | 166 | 37.0 |
| Female | 283 | 63.0 |
| *Highest education level attained* |  |  |
| No formal education | 2 | 0.4 |
| Primary (Approx. 7 years) | 0 | 0.0 |
| O' level (Approx. 11 years) | 26 | 5.8 |
| A' Level (Approx. 13 years) | 11 | 2.5 |
| Tertiary (Colleges, Polytechnic & University) | 410 | 91.3 |
| *Employment status* |  |  |
| Not employed | 37 | 8.2 |
| Informal sector | 51 | 11.4 |
| Formal sector | 332 | 73.9 |
| Students | 29 | 6.5 |
| *Residence* |  |  |
| Urban province | 328 | 73.1 |
| Rural province | 121 | 26.9 |
| *Lockdown support from government, NGOs, religious groups and or private sector* |  |  |
| Yes | 20 | 4.4 |
| No | 429 | 95.6 |
| *If Yes, Specify:* |  |  |
| Financial assistance | 9 | 2.0 |
| Food handouts | 11 | 2.5 |
| Grocery voucher | 1 | 0.2 |
| Not Applicable | 428 | 95.3 |
| *No of people staying at home, median and IQR* | 4 (3:6) |  |

**Notes:** IQR = interquartile range, NGOs =Non-Governmental organisations

### Lifestyle-related habits change during the COVID-19 induced lockdown

Majority of the participants (62.9%) indicated that the lockdown resulted in decreased physical activity levels and alcohol consumption (53.3%). While on the contrary greater proportion of the participants reported an increase in smoking (45.9%) and screen time spent on TV/ tablet/ cell phone/ laptop (90.1%). The results show a disturbing trend of decreased physical activity and increased time spent on sedentary activities.

### Lockdown and food access

**Figure 1** shows that the lockdown resulted in glaring increases in food prices. This was reported by 94.8% of the study participants. In addition, 64% reported a decrease in availability of diverse and nutritious foods, whilst 43.9% mentioned that the quality of foods sold in their areas had decreased. These results show a disturbing picture where food security of most households appears to be compromised due to extortionate prices, availability of less nutritious food choices with poor quality (possible safety concerns as well). Implications of lockdown on dietary diversity

**Figure 1:**
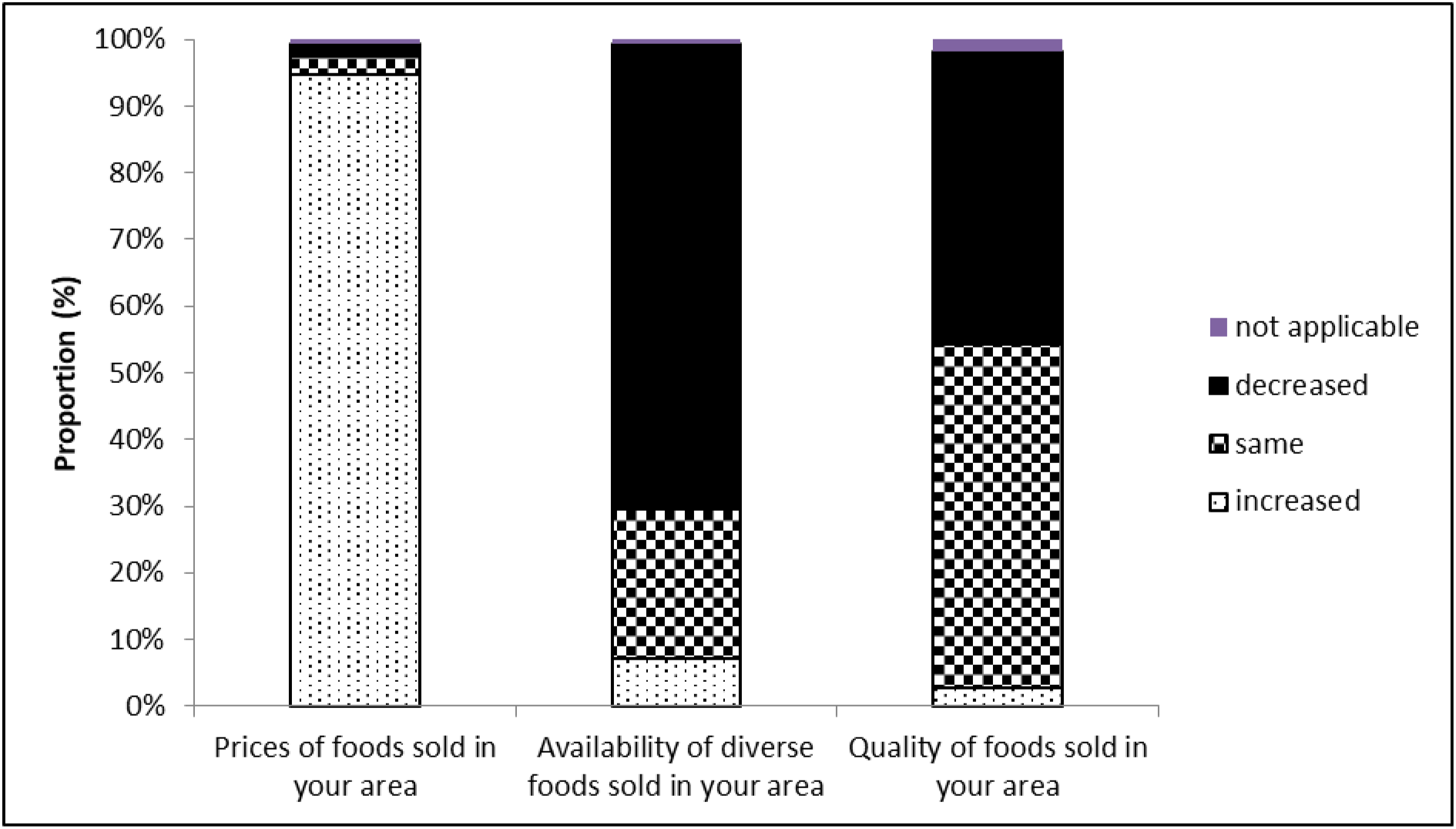
Effect of COVID-19 induced lockdown on food prices, availability and diversity (n=438)

The results show that 96.6% (n=490) of the participants reported that their diet and consumption patterns have changed during the COVID-19 induced lockdown. Concerning individual food groups, 57.8% of the participants stated that there was a decrease in consumption of ‘other vitamin A rich fruits and vegetables’ (**Figure 2**). There was also a decrease in the intake of ‘other vegetables’ (48.5%), ‘other fruits’ (64.9%), ‘nuts and seeds’ (45.0%), ‘cereals breads and tubers’ (41.1%), and ‘dairy products’ (44.9%). Interestingly, we observed an increase in consumption of ‘dark green leafy vegetables’ (33.72%). However, ‘egg’ consumption largely remained the same (41.8%), as well as ‘meat and meat group’ (46.2%). The reported consumption patterns are reflective of a disrupted food system and prevailing food access restrictions during the lockdown.

**Figure 2:**
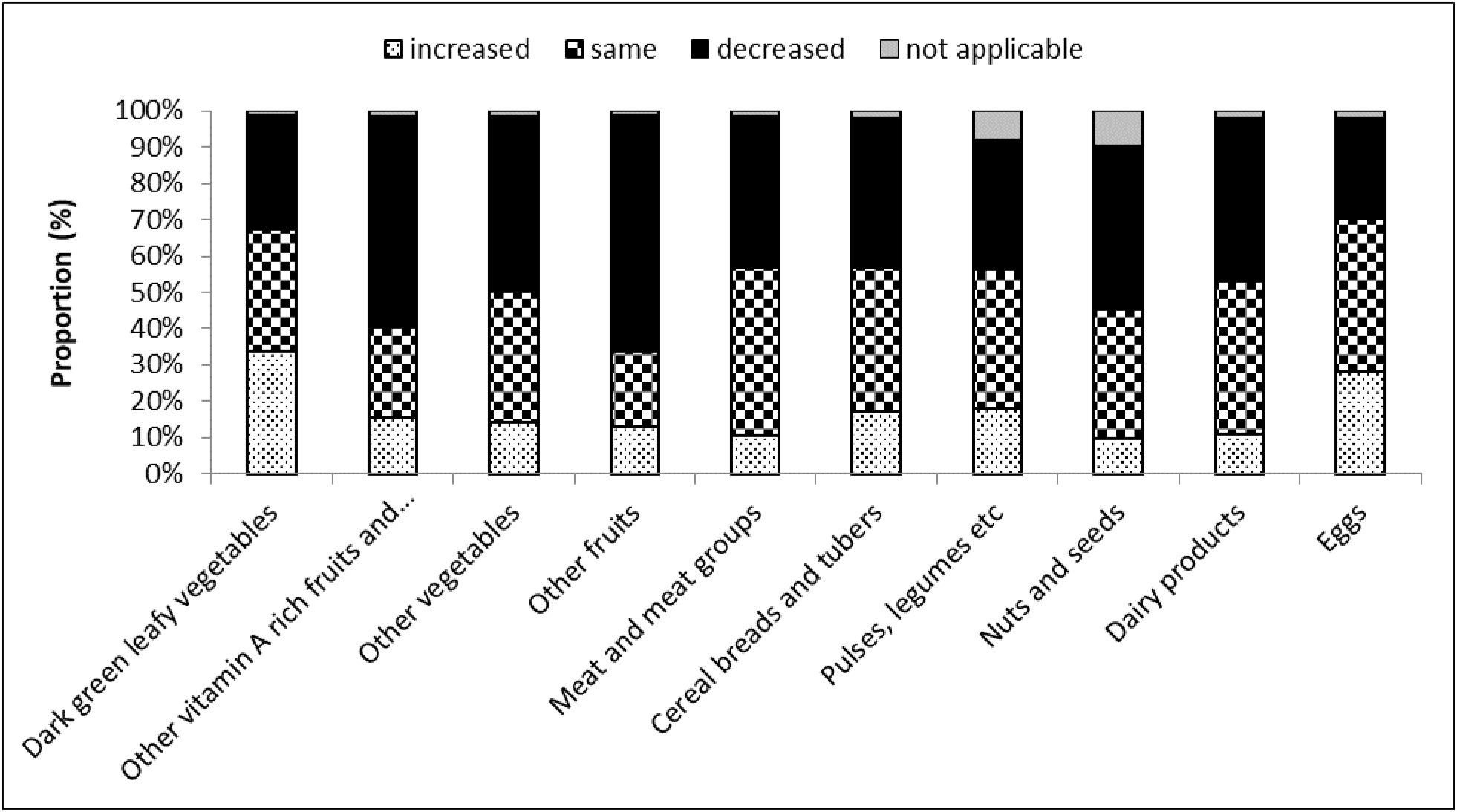
Effect of COVID 19 induced lockdown on consumption of various food groups (n=427)

### Physical activity and lifestyle changes

**Figure 3** shows that most (76.4%) of the participants reported to having increased stress and anxiety, while 89.1% reported increased screen time and 62.5% indicated that the lockdown resulted in reduction in their physical activity levels. Effect of Covid-19 on stress and anxiety levels.

**Figure 3:**
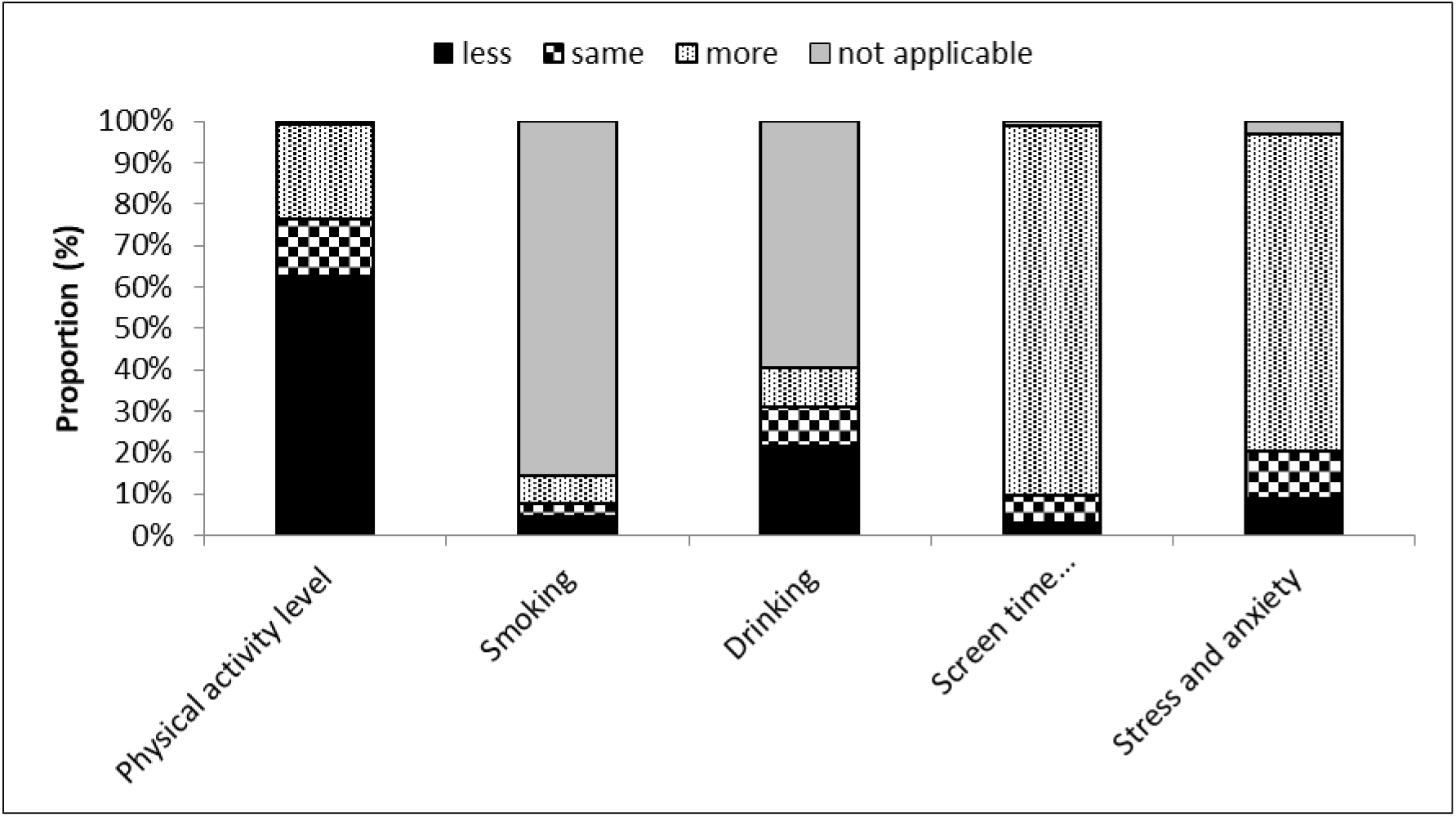
Physical activity and selected lifestyle indicators during the lockdown (n=421)

Concerning stress and anxiety participants mostly reported that they were psychologically affected by the COVID-19 pandemic and associated lockdown restrictions (**Figure 4**). Specifically, 97.1% of the respondents testified that COVID-19 pandemic and associated lockdown had increased their stress and anxiety levels.

**Figure 4:**
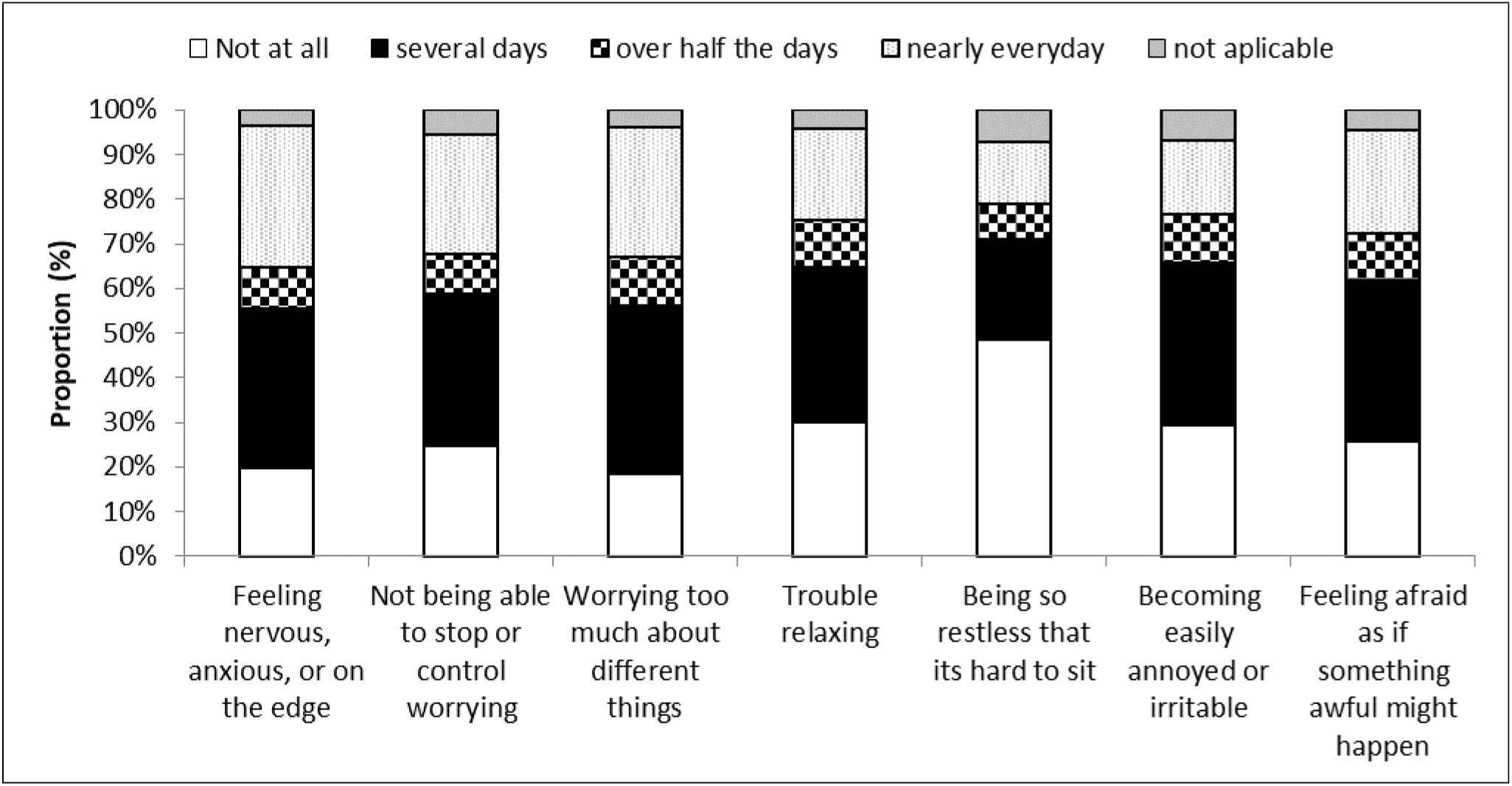
Stress and anxiety during COVID-19 induced lockdown (n=409)

The prevalence of GAD in this study was 40.4% and mostly affected females [63.5%, P=0.909), 31-40 years age group (49.6%, P=0.886) (**Table 2**). The Cronbach’s alpha for the GAD scale in our study was 0.926, which indicates a high level of internal consistency for our scale with this study population. The lockdown had more pronounced impact on reduced physical activity among participants with GAD symptoms (71.5%, P=0.030). While smoking increased (55.2%, P=0.133), alcohol drinking decreased (54%, P=0.898) and screen time (TV, laptop, phone) increased (92.5%, P=0.062) among participants with GAD symptoms.

**Table 2:** GAD and selected participant characteristics

| Variable | Total | GAD symptoms |  | <sup>1</sup>P-value |
| --- | --- | --- | --- | --- |
|  | n (%) | n (%) |  |  |
|  |  | No | Yes |  |
| Age category, years |  |  |  |  |
| 18-30 | 91 (26.8) | 52 (25.7) | 39 (28.5) | 0.886 |
| 31-40 | 167 (49.3) | 99(49.0) | 68 (49.6) |  |
| 41-49 | 47 (13.9) | 30 (14.9) | 17 (12.4) |  |
| 50+ | 34 (10.0) | 21 (10.4) | 13 (9.5) |  |
| Sex |  |  |  |  |
| Male | 126 (37.2) | 76 (37.6) | 50 (36.5) | 0.909 |
| Female | 213 (62.8) | 126 (62.4) | 87 (63.5) |  |
| Highest education level attained |  |  |  |  |
| Primary & below | 16 (4.7) | 8 (4.0) | 8 (5.8) | 0.718 |
| O & A levels | 9 (2.7) | 5 (2.5) | 4 (2.9) |  |
| Tertiary | 314 (92.6) | 189 (93.6) | 125 (91.2) |  |
| Employment status |  |  |  |  |
| Not employed | 29 (8.6) | 18 (8.9) | 11 (8.0) | 0.406 |
| Informal sector | 36 (10.6) | 19 (9.4) | 17 (12.4) |  |
| Formal sector | 252 (74.3) | 155 (76.7) | 97 (70.8) |  |
| Students | 22 (6.5) | 10 (5.0) | 12 (8.8) |  |
| Residence |  |  |  |  |
| Urban province | 254 (74.9) | 162 (80.2) | 92 (67.2) | 0.007* |
| Rural province | 85 (25.1) | 40 (19.8) | 45 (32.8) |  |
| Weight Change (Pre vs Post lockdown) |  |  |  |  |
| Lost weight | 82 (25.3) | 42 (21.8) | 40 (30.5) | 0.143 |
| Gained weight | 148 (45.7) | 89 (46.1) | 59 (45.0) |  |
| Weight did not change | 94 (29.0) | 62 (32.1) | 32 (24.4) |  |

**Notes:** <sup>1</sup>P value based on Pearson's Chi-square test, \*Significant at $P<0.05$ .

The results show that those individuals that exhibited GAD symptoms had reduced intake of vitamin A rich fruits and vegetables (69.1%, P=0.005), other vegetables (58.8%, P=0.001), other fruits (80.1%, P<0.001), meat and meats (54.9%, P=0.001), cereals, bread and tubers (56.7%, P<0.001), pulses and legumes (55.9%, P<0.001), nuts and seeds (60.8%, P=0.006), milk and dairy (60.3%, P<0.001), eggs (42.2%, P<0.001) compared to their less anxious counterparts.

### Body weight and image perceptions

Almost half (48%) thought they had gained weight, whilst 25.8% and 25.5% thought they had lost weight and weight did not change respectively. However, based on the 9 figural BMI-SMT to determine perceived anthropometry 44.5% gained weight, 24.3% lost weight and 31.2% did not have weight change. The average size before lockdown was 5.0±1.6 compared to 5.2±1.8 during lockdown. The paired samples T test showed that there was a significant increase in perceived body size when comparing pre and during lockdown body size (P<0.001). Based on the BMI-SMT we found that highest perceived weight gain during the lockdown was in the age group 31-40 years (48.9%, P=0.568), females (62.6%, P=0.062), participants with tertiary qualifications (92.5%, P=0.533), formal sector employees (75.3%, P=0.107), individuals who did not have GAD symptoms (60.1%, P=0.143).

### Ease of Access to health services

Over half (59.9%) said drugs and medication were not easily available during lockdown compared to 19.8% who said the drugs and medication were still easily available. Over half (58.6%) said doctor’s appointments and review visits were not easily available and 7.97% said appointments and review visits were still easily available. Concerning immunisation and growth monitoring 37.8% said this service was not easily available compared to 19% who mentioned that the services were still easily available. Concerning the same 59.9% this question did not apply to them (**Figure 5**).

**Figure 5:**
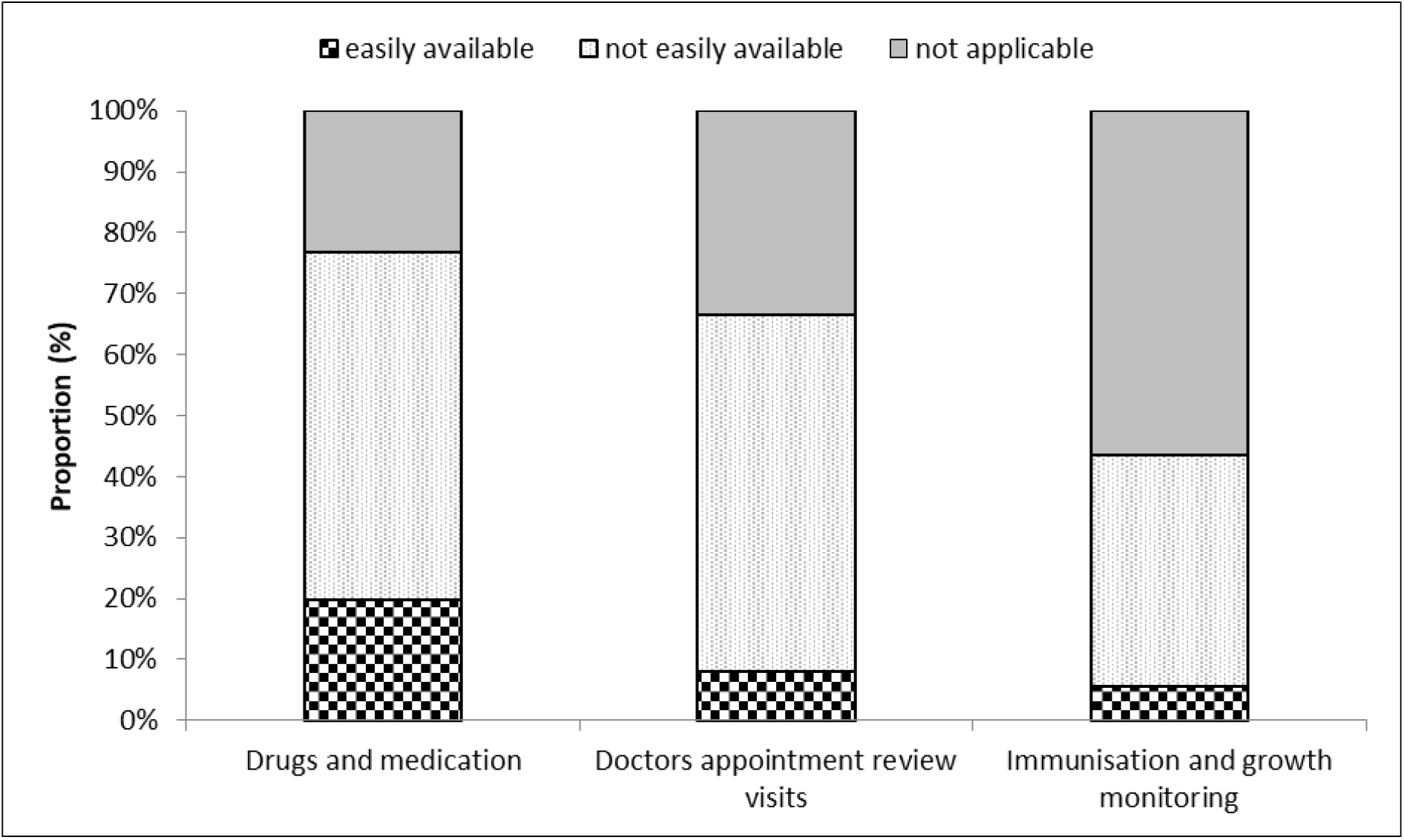
Availability of medication and access to selected health services during lockdown (n=390)

## DISCUSSION

In the present study, we provided for the first-time data on the effect of the COVID-19 lockdown on nutrition, anxiety, physical activity and lifestyle patterns in Zimbabwe. However, there are still lots of unknowns concerning the COVID-19 pandemic that is still ongoing, thus our data need to be confirmed and investigated in future population studies.

The lockdown period (4 weeks) was associated with increases in food prices and decrease in dietary diversification. The increases in food prices is reflective of the negative impact of COVID-19 and lockdowns on agriculture and food supply systems ^25^. This often results in artificial shortages and inflated prices and compromising food security and access to healthy food options particularly on the vulnerable low income households. This is disturbing considering the importance of optimum nutrition for well-functioning immune system in these times of COVID-19 ^26^. Therefore, increases in food prices reduces access to diverse and nutritious foods, which ultimately makes it difficult for individuals to maintain thriving immune systems. Healthy diets help to avoid deficiencies of the nutrients that play an essential role in immune cell triggering, interaction, differentiation, or functional expression ^27^. Considering that there is currently no known cure, the Hippocrates 400BC philosophy of, “let thy food be thy medicine, and let medicine be thy food.” is vindicated as consumption of varied and nutritious foods will guarantee a healthy immune system ^28^.

Unfortunately, the effect of COVID-19 pandemic on nutrition and dietary intake goes beyond the individual and the community to reach national and global levels ^27^. Therefore, there are crucial policy implications for national governments centred around the need to stop the spread of COVID-19 using lockdowns and on the other side the glaring need to prevent and deal with the negative impacts of such policy decisions on food security and livelihoods. According to Siche (2020), the COVID-19 pandemic food demand and thus food security are greatly affected due to mobility restrictions, reduced purchasing power, and with a greater impact on the most vulnerable population groups. It remains the responsibility of the governments to create a supportive policy environment to enhance the physical and mental health of individuals in the context of COVID-19 pandemic, without also neglecting the potential risk of “lockdown associated obesity” during lockdowns. Naja and Hamadeh (2020) gave recommendations on how to provide nutrition demands in the context of COVID-19 utilising a multi-level framework for action adapted from the ecological model of health behaviour. There are economic arguments that prolonged lockdown are not sustainable in the long run, as this will lead to economic slump that ultimately creates negative health consequences “poverty-infection complex”, which create more non–COVID-19-related deaths than confinement would save from this disease ^29^. Governments are encouraged to make evidence and informed decisions to ensure responsible lockdown exit strategies.

The COVID-19 induced lockdown has resulted in and disruptions in consumption patterns (96.6%) and elevated anxiety levels (40.4%) Regarding the implications of lockdown on diet and consumption patterns, participants reported a reduction in intake of fruits and vegetables except for ‘dark green leafy vegetables’. This decrease is disturbing considering that WHO recommends an average consumption of 400 gr of fresh fruits and vegetables per day will boost the immune system. Therefore, the observed increase in consumption of dark leafy vegetables is commendable and reflects the utilisation of home or backyard nutrition gardens, that are common in most households in Zimbabwe ^30^.

In this current study, the prevalence of GAD was 40.4% and mostly affecting females and the 31-40 years age group. Hence the finding that carbohydrate intake also decreased is surprising, considering that we expected this to increase due to the stress and sugar cravings in stressful lockdown conditions ^16^. The current findings reveal that among participants with GAD symptoms, there was reduced physical activity and alcohol intake, while an increase in smoking and screen time (TV, laptop, and phone). In addition, continuously hearing or reading about the COVID-19 can be stressful and can lead towards overeating, mostly sugary foods “*comfort foods*” ^31^.

With respect to physical activity we observed that most of the participants were less active and gained weight in the lockdown period, thus increasing the risk of overweight and obesity. The perceived weight gain by participants was higher in females (P=0.062), participants with tertiary qualifications (P=0.533), formal sector employees (P=0.107). These results paint a picture of the dangers of obesity associated severe COVID-19 complications in typical formally employed persons with a tertiary qualification. Considering that there is growing evidence showing that obesity is key risk factor in this crisis ^8^. It is strongly recommended that individuals should increase physical activity levels and reduce the consumption of energy dense “junk” food which predisposes to weight gain and susceptibility to COVID-19 ^32^. Thus, studies to investigate the impact of consumption of unhealthy diets and low physical activity and anxiety on the susceptibility to COVID-19 and recovery are warranted.

We reported reduction in access to medical doctors (58.6%) said, drugs (59.9%), immunisations and growth monitoring (37.8%) during the lockdown period. This has negative long-term implications as individuals will develop reluctance to access preventative health services. In addition, disruptions in drug supply chains is likely associated defaulters on immunisations schedules among children and this may lead to future fatal outbreaks of preventable diseases such as measles polio and diphtheria ^33^.

### Limitations

The main limitation of the present study is that we utilised an online and self-reported questionnaire with completion rate of 76%, thus missing values on some variables. Although our survey had respondents from all the 10 provinces of Zimbabwe, online surveys tend to be restricted to individuals with access to internet with potential under representation of people from lower socio-economic groups and rural settings. However, due to the lockdown travel restrictions the online survey was the only option to collect this critical dietary and physical activity data. Although the BMI-based Silhouette Matching Test (BMI-SMT) has its limitations with respect to collective objective anthropometry data, in the current lockdown context it remains useful alternative. It’s crucial that this rapid assessment went on at the most critical period of the COVID-19 pandemic, to inform program and policy decisions. Interestingly, evidence is gathering indicating that web based surveys are equivalent to conventional face to face interviews ^34,35^. There is growing concern that COVID-19 could deepen food insecurity, malnutrition in Africa. Unfortunately, in our current study we did not collect data to understand the different food groups and how they are affected by the hike in prices, this may have helped to clearly understand the impact of lockdown on food access and household food security issues.

## CONCLUSIONS

The lockdown period was associated with increases in food prices and decrease in dietary diversification. In addition, the COVID-19 induced lockdown has resulted in disrupted consumption patterns and elevated anxiety levels. Participants indicated that their physical activity levels decreased and perceived weight gain in the lockdown period, thus increasing the risk of overweight and obesity. However, there are still lots on unknowns concerning the COVID-19 pandemic, as such future studies with incorporating participants of different socio-economic status are warranted.

## Data Availability

The anonymised datasets used and/or analysed during the current study are available from the corresponding author on reasonable request. Please for data requests.

## Acknowledgements

Firstly, we are very grateful to the respondents for accepting to join our rapid appraisal and for sharing their perspectives and insights into perceived effects of the COVID-19 in their communities. Great thank you to Ms Tavonga Muderedzwa for assistance in setting up the online poll.

## Contributors

All authors participated in the design and conceptualisation of the project, data analysis and interpretation and writing of the manuscript. All authors read and approved the final manuscript.

## Patient consent for publication

Not required.

## Consent for publication

No previously published material has been used in this study.

